# Diagnosis of COVID-19 using CT scan images and deep learning techniques

**DOI:** 10.1101/2020.07.11.20151332

**Authors:** Vruddhi Shah, Rinkal Keniya, Akanksha Shridharani, Manav Punjabi, Jainam Shah, Ninad Mehendale

## Abstract

Early diagnosis of the coronavirus disease in 2019 (COVID-19) is essential for controlling this pandemic. COVID-19 has been spreading rapidly all over the world. There is no vaccine available for this virus yet. Fast and accurate COVID-19 screening is possible using computed tomography (CT) scan images. The deep learning techniques used in the proposed method was based on a convolutional neural network (CNN). Our manuscript focuses on differentiating the CT scan images of COVID-19 and non-COVID 19 CT using different deep learning techniques. A self developed model named CTnet-10 was designed for the COVID-19 diagnosis, having an accuracy of 82.1 %. Also, other models that we tested are DenseNet-169, VGG-16, ResNet-50, InceptionV3, and VGG-19. The VGG-19 proved to be superior with an accuracy of 94.52 % as compared to all other deep learning models. Automated diagnosis of COVID-19 from the CT scan pictures can be used by the doctors as a quick and efficient method for COVID-19 screening.

## 1 Introduction

The new Coronavirus infection was first reported in Wuhan, China, and since then it has strongly spread out since January 2020 worldwide. The World Health Organization (WHO) declared the outbreak from the Corona virus disease 2019 (COVID-19) to be a public health emergency of international concern on the 30th of January, 2 020. The specific symptoms of COVID-19 are fever, dry cough, sore throat, loss of taste, or smell. In addition to this, the unspecific symptoms are tiredness, headaches, and breathlessness (3 %). Normally, it takes 5-6 days for the symptoms to be shown in the patient’s body. Animals are also able to transmit this infection and themselves getting affected. Mainly two similar viruses were reported earlier, which were Severe Acute Respiratory Syndrome Coronavirus (SARS-CoV) and the Middle East Respiratory Syndrome Coronavirus (MERS Coronavirus). These viruses caused major respiratory problems and are zoonotic in nature.

In the current scenario, the COVID-19 test results take more than 24 hours to detect the virus in the human body. There is an urgent need to recognize the illness in the early stage and to put the infected immediately under quarantine because no specific drugs are available for COVID-19. The Chinese government reported that the diagnosis is confirmation of COVID-19 by the real-time polymerase chain reaction (RT-PCR). RT-PCR suffers from high false-negative rates and utilizes a lot of time. The low sensitivity RT-PCR test is not satisfactory in the present pandemic situation. In some cases, the infected are not possibly recognized on time and do not receive suitable treatment. The infected can be assigned sometimes as COVID-19 to healthy people because of a false-negative result.

In comparison to RT-PCR, the Thorax Computer Tomography (CT) is possibly more reliable, useful, and quicker technology for the classification and assessment of COVID-19, in particular to the epidemic region. Almost all hospitals have of CT-Image Screening; hence the Thorax CT pictures can be used for the early detection of COVID-19 patients. Nevertheless, the COVID-19 classification basing on the Thorax CT requires a radiology expert, and a lot of valuable time is lost. Hence, automated analysis of the Thorax CT pictures is desirable to save the valuable time of the medical specialist staff. This will also avoid delays in starting treatment.

Deep Learning is the most efficient technique that can be used in medical science. It is a fast and efficient method for the diagnosis and prognosis of various illnesses with a good accuracy rate. They are specifically trained models to classify the inputs into different categories desired by the programmers. In the medical field, they are used to detect heart problems, tumors using image analysis, diagnosing cancer, and many other applications. It is also used to differentiate the CT Scan images of the patients infected with COVID-19 as positive or not infected i.e. negative. A self-developed model CTnet-10 was created having an accuracy of 82.1 %. To improve the accuracy we had also passed the CT scan image through multiple preexisting models. We found that the VGG-19 model is best to classify the images as COVID-19 positive or negative as it gave a better accuracy of 94.52 % accuracy. A graphical representation of our proposed model is demonstrated in Figure 1. The CT scan image is passed through a VGG-19 model that categorizes the CT scan into COVID-19 positive or COVID-19 negative.

**Fig. 1:**
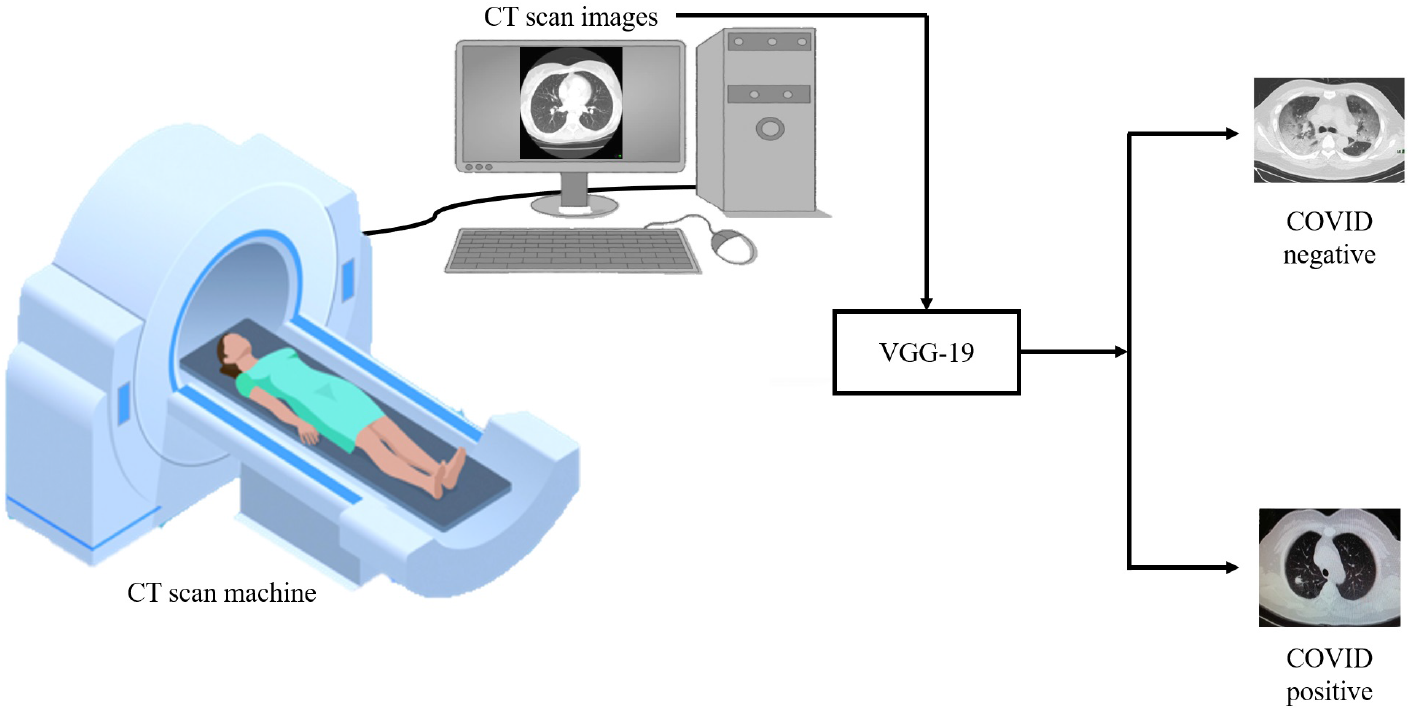
System flow diagram. The CT scan machine gives the CT scan image of a patient for the screening of COVID-19. The input image is passed through a VGG-19 model that categorizes the image as COVID-19 positive or negative.

## 2. Literature review

Several studies and research work have been carried out in the field of diagnosis from medical images such as computed tomography (CT) scans using artificial intelligence and deep learning. DenseNet architecture and recurrent neural network layer were incorporated for the analysis of 77 brain CTs by Grewal *et al*. [1]. RADnet demonstrates 81.82 % hemorrhage prediction accuracy at the CT level. Three types of deep neural networks (CNN, DNN, and SAE) were designed for lung cancer calcification by Song *et al*. [2]. The CNN model was found to have better accuracy as compared to the other models. Using deep learning, specifically convolutional neural network (CNN) analysis, Gonzalez *et al*. [3] could detect and stage chronic obstructive pulmonary disease (COPD) and predict acute respiratory disease (ARD) events and mortality in smokers.

During the outbreak time of COVID-19, CT was found to be useful for diagnosing COVID-19 patients. The key point that can be visualized from the CT scan images for the detection of COVID-19, was ground-glass opacities, consolidation, reticular pattern, and crazy paving pattern [4]. A study was done by Zhao *et al*. [5] to investigate the relation between chest CT findings and the clinical conditions of COVID-19 pneumonia. Data on 101 cases of COVID-19 pneumonia were collected from four institutions in Hunan, China. Basic clinical characteristics and detailed imaging features were evaluated and compared. A study on the chest CTs of 121 symptomatic patients infected with coronavirus was done by Bernheim *et al*. [6]. The hallmarks of COVID-19 infection as seen on the CT scan images were bilateral and peripheral ground-glass and consolidative pulmonary opacities. As it is difficult to obtain the datasets related to COVID-19, an open-sourced dataset COVID-CT, which contains 349 COVID-19 CT images from 216 patients and 463 non-COVID-19 CTs was built by Zhao *et al*. [7]. Using the dataset, they developed an AI-based diagnosis model for the diagnosis of COVID-19 from the CT images. On a testing set of 157 international patients, an AI-based automated CT image analysis tools for detection, quantification, and tracking of coronavirus was designed by Gozes *et al*. [8]. The accuracy of the model developed was 95 %. The common chest CT findings of COVID-19 are multiple ground-glass opacity, consolidation, and interlobular septal thickening in both lungs, which are mostly distributed under the pleura [9]. A deep learning-based software system for automatic COVID-19 detection on chest CT was developed by Zheng *et al*. [10] using 3D CT volumes to detect COVID-19. A pre-trained UNet and a 3D deep neural network were used to predict the probability of COVID-19 infections on a set of 630 CT scans. Of 1014 patients, 601 patients tested positive for COVID-19 based on RT-PCR and the results were compared with the chest CT. The sensitivity of chest CT in suggesting COVID-19 was 97 % as shown by Ai *et al*. [11]. In a series of 51 patients with chest CT and RT-PCR tests performed within 3 days by Fang *et al*. [12], the sensitivity of CT for COVID-19 infection was 98 % compared to RT-PCR sensitivity of 71 %. An AI system (CAD4COVID-Xray) was trained on 24,678 CXR images including 1,540 used only for validation while training. The radiographs were independently analyzed by six readers and by the AI system. Using RT-PCR test results as the reference standard, the AI system correctly classified CXR images as COVID-19 pneumonia with an AUC of 0.81 [13].

## 3 Methodology

The images were collected from the COVID-19 CT dataset. A total of 738 images were split into a training set, validation set, and test set with a split of 80 %, 10 %, 10 % respectively. The COVID-19 CT dataset had 349 CT images containing clinical findings of COVID-19 from 216 patients. It was used to classify a slice of CT scan images to COVID-19 and non-COVID-19. We trained CTnet-10, the input size was 128×128×3. Both the convolutional block I and II consisted of 2 layers and a pooling layer with a flatting layer to convert 2D to 1D layer, which was fed to a dense layer of 256 neurons. Dropout of value 0.3 had been added in which the output layer contained 1 neuron and then the result had been predicted to classify the CT scan images. The self-developed network (CTnet-10) was fed with an input image of 128×128×3 and at every layer the dimensions of the activations changes as shown in the Figure 2. Both the convolutional block I and II consisted of 2 layers and a pooling layer with a flatting layer to covert 2D to 1D layer, which was fed to a dense layer of 256 neurons. Dropout of value 0.3 had been added in which the output layer contained 1 neuron and then the result had been predicted to classify the CT scan images.

**Fig. 2:**
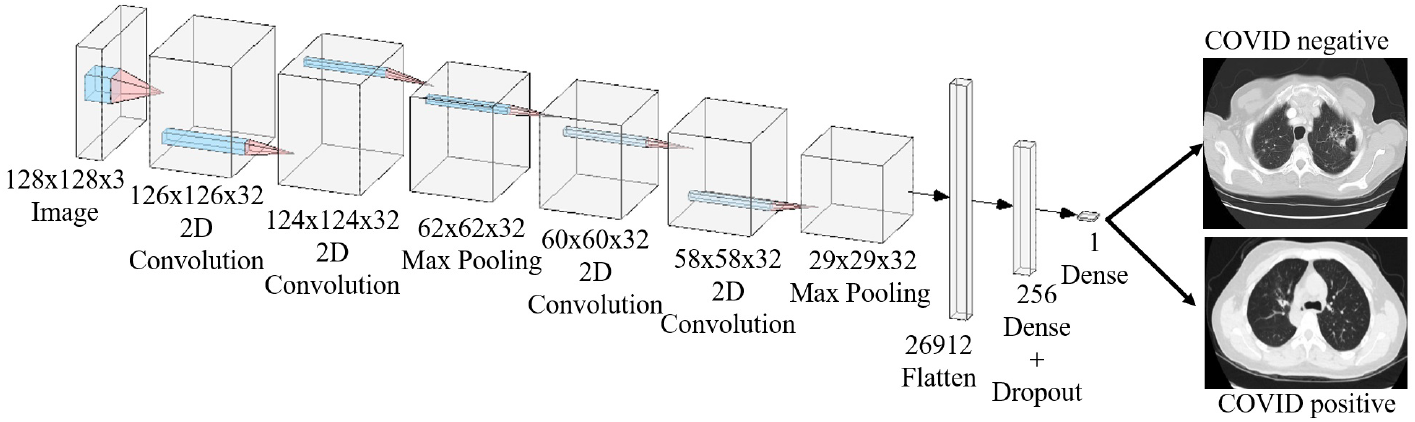
Configuration of the CTnet-10 model. The model was fed with an input image of size 128×128×3. There are a total of 4 convolutional blocks. It passes through 2 convolutional blocks of dimensions 126×126×32, 124×124×32 respectively. Then it passes through a max-pooling of dimension 62×62×32 followed by 2 convolutional layers of dimensions 60×60×32, 58×58×32 respectively. This is further passed through a pooling layer of dimension 29×29×32. It is then passed through 26912 neurons of the flattened layer, which is further passed through dense and dropout layers of 256 neurons each. After passing it through a single neuron of a dense layer, the CT scan images are classified as COVID-19 positive or negative.

For the VGG-19 model, the image resolution used was (224, 224, 3). The VGG-19 network was fed with an input image of 224×224×3 and it outputs a number between 0 and 1. For this case, less than 0.5 corresponds to COVID-19 positive and greater than or equal to 0.5 implies COVID-19 negative. As mentioned above we used VGG-19 architecture with pre-trained weights of imagenet. It is a 24-layer model which consists a total of 5 convolutional blocks, 3 max pool layers and 3 FC layers, but we did a fine-tuning by using pre-trained weights for all convolutional blocks, removing the last two fully connected (FC) layer and then adding 2 FC layer with 4096 neurons, dropout was used with each of these layers for regularization with a rate of 0.3. The final binary classification layer of single-neuron governed by sigmoid activation was added. The model was compiled with ADAM optimization with the default learning rate; the loss function used was binary cross entropy. The model was trained on a batch size of 32 and EarlyStoping was used to prevent overfitting. First, it was trained on 30 epochs without EarlyStoping, and then on 20 epochs with EarlyStoping, the model stopped at epoch no. 10. The VGG-19 network was fed with an input image of 224×224×3 and at every layer, the dimensions of the activations change as shown in the Figure 3. It was a 24-layer model that consists of a total of 5 convolutional blocks, 3 max pool layers and 3 FC layers, but we did a fine-tuning by using pre-trained weights for all convolutional blocks, removing the last two FC layer and then adding 2 FC layer with 4096 neurons. Dropout was used with each of these layers for regularization with a rate of 0.3. After the image has gone through all the layers just described we are left with a scalar between 0 and 1 as our final output and if it’s greater than 0.5 then it corresponds to COVID-19 negative and less than it corresponds to COVID-19 positive.

**Fig. 3:**
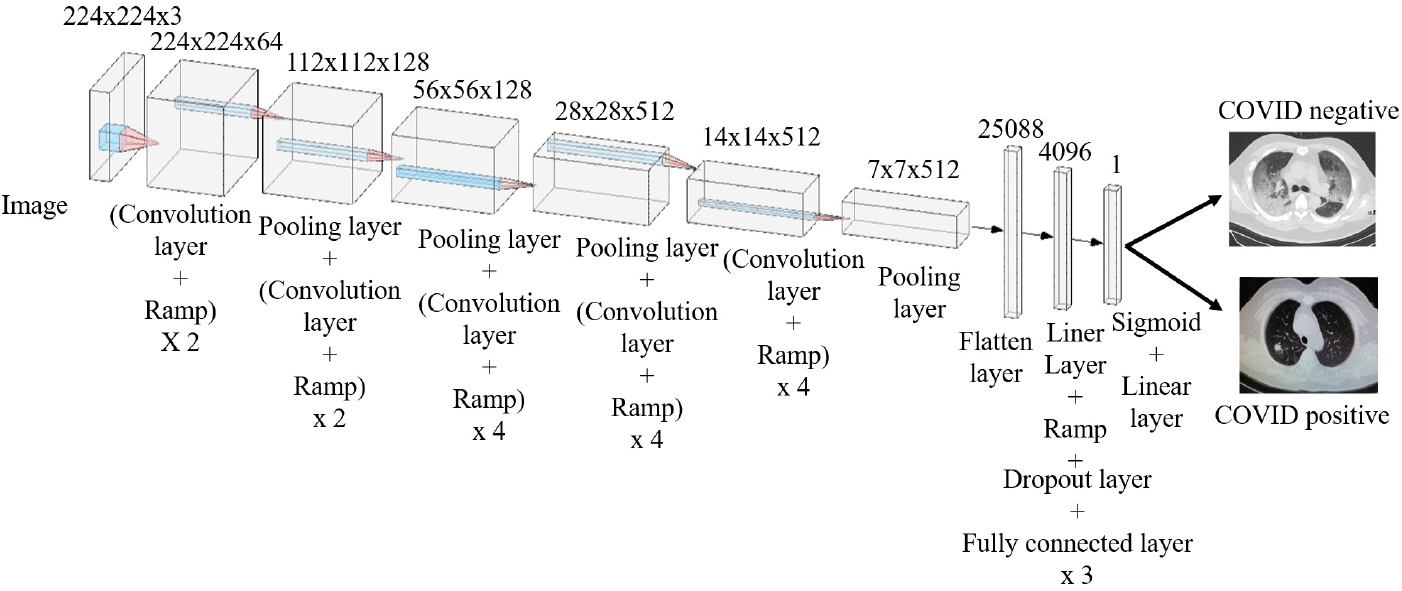
Configuration of the model of VGG-19. The model was fed with an input image of size 224×224×3. There are a total of 5 convolutional blocks. It passes through one of the convolutional layers and a ramp of dimension 224×224×64. Then it passes through a next pooling and convolutional layer of dimension 112×112×128. Then it again passes through two pooling layers of dimensions 56×56×128, 28×28×512 respectively. This is further passed through the consecutive layer of dimensions 14×14×512, and a pooling layer of 7×7×512. It is then passed through 25088 neurons of the flattened layer, which is consecutively passed through an FC layer of 4096 neurons, in which the dropout layer was used in each of these. After passing it through a single neuron sigmoid and linear, the CT scan images are classified as COVID-19 positive or negative.

The input images were fed to the visual geometry group-16 (VGG-16) model with a dimension of 150×150×3. The model consists of 19 layers, having 5 convolutional blocks each block consisting of two or three convolutional layers and 5 max-pooling layers, finally ending with 2 fully connected (FC) and a softmax layer. We just replaced the softmax layer with the sigmoid layer for binary classification purposes. The model was trained with root mean square propagation (RMSPROP) and a learning rare (2* *e*^*−* 5^) for 30 epochs. We also tried image augmentation on the same model and trained it for 100 epochs.

For the Inception V3 model, the image resolution was kept at 224×224×3. 2 dense layers were added of 1000 and 500 neurons with an L1 and L2 regularization respectively, which were set to 0.01. The ResNet-50 model’s resolution was also kept at 224×224×3. It had 50 layers of dropout. The top layer was not included in this model.

Then we tried the DenseNet-169 model. It is a state of art model which has 169 layers in it. The learning rate used was (1\**e*^*−* 4^) with RMSPROP and the model was trained for 30 epochs. The size of the images was kept at 224×224×3, and a total of 73 images were used for this model. All the deep learning codes, are provided in the Supplementary information.

## 4 Results and discussions

To validate the results, we first trained the CTnet-10 model network using 592 labeled images via supervised learning with 74 validation images. The accuracy of CTnet-10 model was 82.1 %. 73 images were used to test the pre-trained VGG16 network which resulted in an accuracy of 89 % between COVID-19 infected and non-infected CT scans. Using Image Augmentation and fine-tuning (by making the layers ‘block5 conv1’, ‘block4 conv1’ trainable), we could get an accuracy of up to 93.15 %. For the Inception V3 model, we achieved 53.4 % accuracy. Now, we used the ResNet model, which did not contain a top layer. The accuracy achieved was 60 %. To improve this, the images were used to train the DenseNet-169 Network, and the ratio of training, validation, and testing were 80:10:10., with 93.15 % accuracy. Now, 73 images were used to test the model having 2 Conv blocks, 2 pooling, 2 FC layers, which resulted in an accuracy of 84 % between COVID-19 positive and negative CT Scans. Then we realized that we have very little data to create our models so we used a VGG-19 pre-trained network for transfer learning and that resulted in 91.78 % accuracy. Then doing some fine-tuning mentioned in the methodology and Image augmentation we were able to reach up to an accuracy of 94.52 % which was by far the best of all other models we tried out.

Figure 4 (a) shows the confusion matrix of model VGG-19, out of 34 COVID-19 positive CT scans images, 32 images were classified as COVID-19, and 2 were wrongly classified as non-COVID-19. In the case of the 39 non-COVID-19 images, 37 were classified as non-COVID-19 and 2 were classified as COVID-19. Figure 4 (b) depicts the confusion matrix for CTnet-10, out of 35 images for COVID-19 positive CT scans, 28 were rightly classified, and 7 were classified as non-COVID-19. 38 images of non-COVID-19 were used, of which 32 were classified non-COVID-19 and 2 were wrongly classified. Figure 4 (c) shows the confusion matrix of VGG-16, 30 images out of 34 were correctly classified as COVID-19, and 4 were wrongly classified whereas for non-COVID-19, out of 39 images 4 were wrongly classified as COVID-19. In the case of the DenseNet-169 network as shown in Figure 4 (d), 31 images were correctly classified as COVID-19 out of 34 images. Out of 39 non-COVID-19 images 37 were classified as non-COVID-19 and 2 were wrongly classified as COVID-19. Figure 4 (e) shows the ResNet-50 model. The confusion matrix states that out of 34 images of COVID-19, 29 were classified as COVID-19, and 5 were wrongly classified. In the case of non-COVID-19 images, all 39 images were correctly classified as non-COVID-19. Figure 4(f) shows the confusion matrix of Inception V3, all 34 COVID-19 images were wrongly classified as non-COVID-19 and 39 non-COVID-19 images were correctly classified as non-COVID-19.

**Fig. 4:**
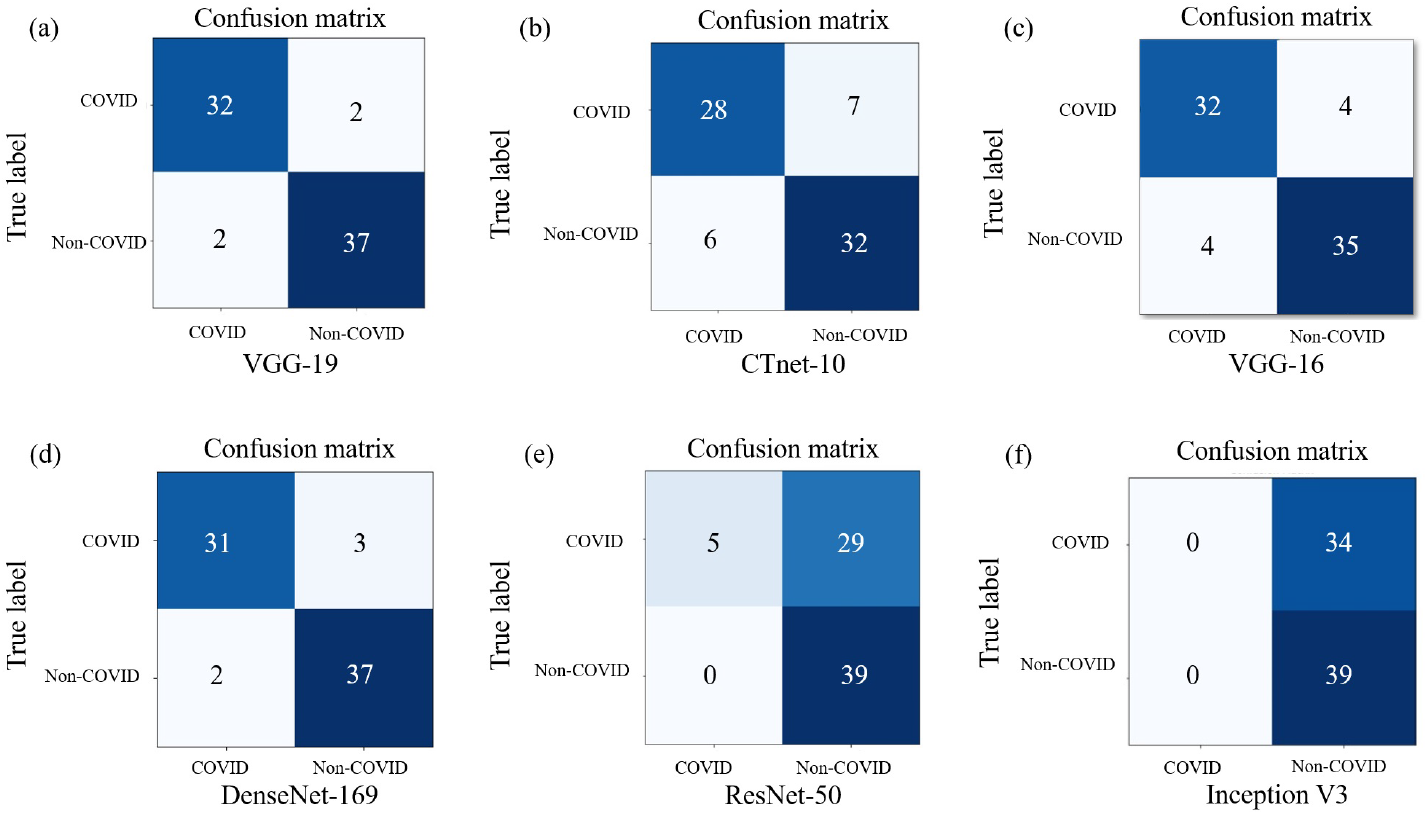
Confusion matrix of the different deep learning models used. (a) Confusion matrix of VGG-19: Out of 34 COVID-19 positive CT scans, 32 were classified as COVID-19 whereas 2 were wrongly classified as Non-COVID-19. Out of 39 non-COVID-19 CT scans, 37 were correctly labeled as non-COVID-19, and 2 were wrongly classified. (b) Confusion matrix of the Self-developed model. 28 out of 35 images were correctly classified as COVID-19. Out of 38 images, 32 were correctly labeled as non-COVID-19 whereas 6 were labeled as COVID-19. (c) Confusion matrix of VGG-16. Out of 34 CT scans for COVID-19, 30 were correctly labeled as COVID-19 whereas 4 were wrongly classified. 35 out of 39 CT scans were correctly labeled as non-COVID-19 and 4 were labeled as COVID-19. (d) Confusion matrix for DenseNet-169. 31 images was correctly labeled as COVID-19 and 3 were wrongly classified as non-COVID-19. Out of 39 non-COVID-19 CT scans, 37 were correctly labeled as non-COVID-19. (e) Confusion matrix for ResNet-50. 29 images were correctly classified as COVID-19 whereas 5 were wrongly classified. 39 out of 39 images were correctly classified as non-COVID-19. (e) Confusion matrix of InceptionV3. 34 out of 34 images were wrongly classified as COVID-19 and 39 images were correctly labeled as non-COVID-19.

The graph shown in Figure 5 is a comparison between accuracy in percentage for 6 different deep learning networks that have been used in our study. The VGG-19 network got the highest accuracy of 94.52 %. The next highest accuracy of 93.15 % was seen in a DenseNet-169 network. The VGG-16 model has an accuracy of 89 %. Our self-developed model achieved an accuracy of 82.1 %, more than the ResNet-50 network which has an accuracy of 60 % and InceptionV3 with an accuracy of 53.4 %.

**Fig. 5:**
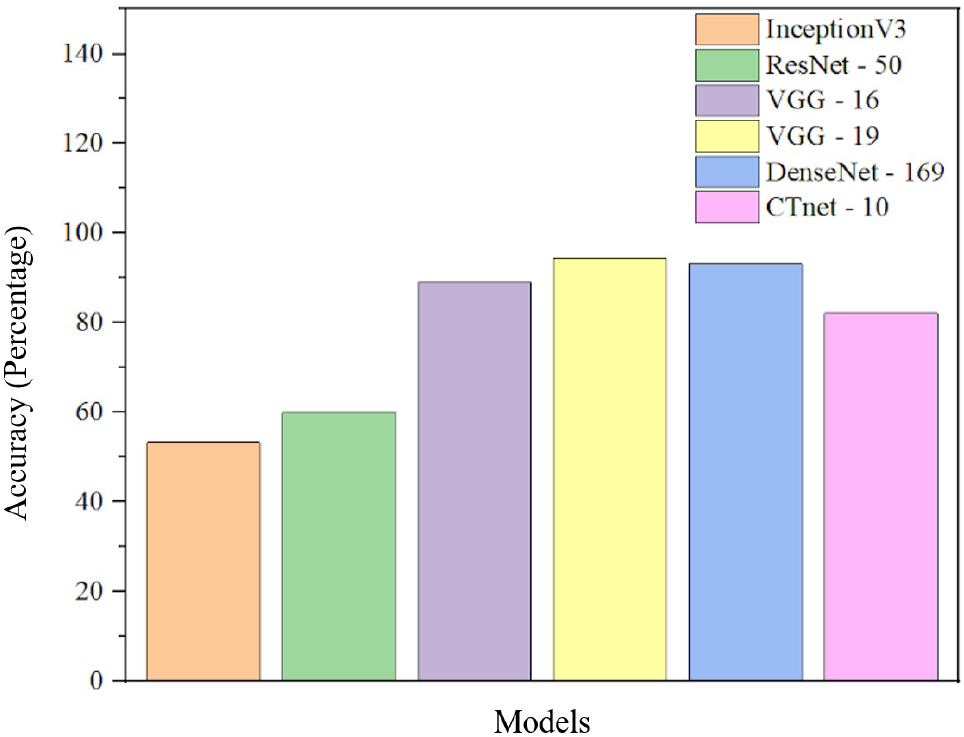
Comparison of accuracy vs. different deep learning networks. VGG-19 has the highest accuracy amongst the different models used in our work. DenseNet-169 has the second-highest accuracy of 93.15 %. The least accurate model was InceptionV3. VGG-16 model shows an accuracy of 89 %. Our self-developed model shows a better accuracy of 82.1 % as compared to ResNet-50 with 60 % accuracy and InceptionV3.

As shown in table 1, we have analyzed 738 CT scan images of the patients available as an open-source data set which comparable number to the reported literature Ai *et al*. [11]. Fang *et al*. [12] has achieved the highest accuracy of 98 %, but they have less number of samples. It is then followed by Ai *et al*. [11] and Gozes *et al*. [8] with an accuracy of 97 % (1014 samples) and 95 % (157 samples) respectively. Our proposed method using the VGG-19 (Proposed method - 2) model gave us an accuracy of 94.52 % and CTnet-10 model (Proposed method - 1) with and accuracy of 82.1 % using 738 CT scan samples. The accuracy of CTnet-10 can be improved further by optimization and fine-tuning.

**Table 1:** Comparison with other methods. All the above models classify the CT scan images as COVID-19 positive or negative with different accuracies according to the specific models

| Method | Number of samples | Accuracy (%) |
| --- | --- | --- |
| Zhao *et al.* [5] | 101 | 78.6 |
| Bernheim *et al.* [6] | 121 | 88 |
| Gozes *et al.* [8] | 157 | 95 |
| Zheng *et al.* [10] | 630 | 90.1 |
| Ai *et al.* [11] | 1014 | 97 |
| Fang *et al.* [12] | 51 | 98 |
| Ctnet-10 (Proposed method - 1) | 738 | 82.1 |
| VGG-19 (Proposed method - 2) | 738 | 94.5 |

CT Scan images can be used for the COVID-19 screening of patients. It gives a detailed image of the particular area, using which we can detect the internal defects, injuries, dimensions of the parts, tumors, etc. Compared to the current RT-PCR method, a CT scan is a reliable method. It is an efficient method for the classification of the images of COVID-19 patients. The results are provided accurately and quickly. There are some side effects of CT scan screening that patients can get exposed to radiation if multiple CT scans are conducted.

## 5 Conclusion

Convolutional neural network (CNN) is quite an efficient deep learning algorithm in the medical field since we get an output just by processing the CT scan images to the respective model. The CTnet-10 model had very well classified the images as COVID-19 positive or negative. Our other models have provided us with much better accuracy. The method used by us is well-organized one that can be used by the doctors for the mass screening of the patients. It will yield better accuracy and at a faster rate as compared to the current RT-PCR method. With the above method for the classification of the CT scan images of the COVID-19 patients, data can be extracted, which would help the doctors to get the information feasibly and quickly.

## Data Availability

Data can be made available

## acknowledgements

The authors would like to thank Ninad’s research Lab for providing image dataset.

## Compliance with Ethical Standards

### Involvement of human participant and animals

This article does not contain any studies with animals or Humans performed by any of the authors. All the necessary permissions were obtained from the Institute Ethical Committee and concerned authorities.

### Information about informed consent

No informed consent was required as the studies does not involve any human participant.

### Funding information

No funding was involved in the present work.

